# Intestinal Parasites among Primary School Children in Aden, Yemen

**DOI:** 10.1101/2024.08.12.24311851

**Authors:** Ali N. M. Gubran, Naif Mohammed Al-Haidary, Marwa Faisal M. Bajubair, Afrah Mohsen Ali Algibary, Manal Galeb Mhmad Ali, Marwa Fuad Othman Ali

## Abstract

**Background:** Intestinal parasite infection is a significant public health problem worldwide. This study aimed to determine the prevalence of intestinal parasites among primary school children, identify the most common types of parasites, and identify the risk factors contributing to infection in Aden, Yemen.

**Methodology/Principal Findings:** An analytical cross-sectional study was conducted on 201 school children in Aden, Yemen. Stool specimens were collected and tested using direct methods (saline and iodine preparations) and sedimentation concentration techniques. Data analysis was performed using SPSS (Version 21) with p ≤ 0.05 considered statistically significant. The overall prevalence of intestinal parasites was 47.3%; 35.8% had a single parasite and 11.5% had multiple parasites. Higher rates were observed among female schoolchildren (51.2%), children whose mothers had primary education (51.3%), secondary education (50%), housewives (48.5%), and children aged >9 years (50%). The most predominant parasite was Entamoeba histolytica/dispar (36.3%). There was no significant association between the identified risk factors and intestinal parasitic infections.

**Conclusions/Significance:** The prevalence rate of intestinal parasites is high in Aden, Yemen, with Entamoeba histolytica/dispar being the most dominant parasite. The highest rates were found among female schoolchildren, those whose mothers were housewives with primary or secondary education, and children aged >9 years.

**Author Summary:** Intestinal parasite infections are a major health issue in many parts of the world, especially in developing countries like Yemen. This study investigated the prevalence of these infections among primary school children in Aden, Yemen. By analyzing stool samples from 201 children, we found that nearly half were infected with at least one type of parasite. The most common parasite was Entamoeba histolytica/dispar, affecting more than a third of the children. We also discovered that certain groups of children were more likely to be infected, such as older children, girls, and those whose mothers had lower levels of education or were housewives. Understanding these patterns can help in developing targeted interventions to reduce the spread of these infections and improve children’s health and educational outcomes.

## Introduction

Neglected tropical diseases (NTDs) represent a diverse group of communicable diseases that prevail in tropical and subtropical conditions in 149 countries, affecting more than one billion people. Among these, parasitic infections remain a significant public health challenge, particularly in developing countries where they are often overlooked by public health agendas. Children are especially vulnerable to these infections due to their developing immune systems, poor hygiene practices, and frequent contact with contaminated soil and water [1,2].

Intestinal parasitic infections, including soil-transmitted helminths (STH) and protozoa, are widespread in regions with inadequate sanitation and limited access to clean water. These infections can lead to severe health outcomes, including malnutrition, impaired physical and cognitive development, and increased susceptibility to other infections. The World Health Organization (WHO) estimates that more than 267 million preschool-age children and over 568 million school-age children live in areas where these parasites are intensively transmitted [3].

Despite their significant impact, parasitic infections are often neglected in terms of research funding, policy prioritization, and healthcare resources. This neglect is due in part to their association with poverty and the marginalized populations they predominantly affect. Consequently, there is a pressing need for comprehensive epidemiological data to inform targeted interventions and control strategies [4].

In Yemen, intestinal parasitic infections among children remain a critical public health issue. The country’s ongoing conflict and resulting humanitarian crisis have exacerbated the conditions that facilitate the transmission of these infections, including displacement, overcrowding, and deteriorating sanitation infrastructure. Understanding the prevalence and risk factors associated with intestinal parasitic infections in this context is essential for developing effective public health interventions [5,6].

Different studies have reported varying prevalence rates of intestinal parasitic infections among children, such as 90% in Al-Mahweet, 51.26% and 27.8% in Taiz, 57.4% in Ibb, and 59% in Hadhramout [7-10]. However, the prevalence among schoolchildren in the Aden governorate remains unclear. This study aims to determine the prevalence of intestinal parasites among primary school children, identify the most common types of parasites, and identify the risk factors contributing to intestinal parasite infection among these children in Aden, Yemen.

By conducting this study, we aim to highlight the burden and determinants of intestinal parasitic infections in a neglected tropical disease-endemic region, thereby advocating for enhanced efforts to address this critical public health challenge.

## Materials and Methods

### Study Design

An analytical cross-sectional study was conducted on 201 primary school children in Aden, Yemen, between November 2023 and April 2024. The study included children aged 7 to 15 years who were randomly selected from 5 primary schools.

### Study Population and Setting

The study included children aged 7 to 15 years who were randomly selected from 5 primary schools in Aden, Yemen. Inclusion criteria were all primary school children within the specified age range who provided stool samples and completed questionnaires. Exclusion criteria were children with chronic illnesses or those who did not provide consent.

### Data Collection

Data were collected using a predesigned and pilot-tested questionnaire. The questionnaire included information on the children’s age, sex, education level, and occupation of their mothers, as well as questions about risk factors contributing to the prevalence of intestinal parasites. The questionnaire was administered by trained field workers.

### Ethical Considerations

Ethical approval was obtained from the Ethics Committee of the University of Science and Technology, Aden, Yemen (MEC No. MEC/AD022). Written informed consent was obtained from the parents or guardians of all participating children.

### Sample Collection and Testing

Stool specimens were collected from all participating children using sterile containers. These specimens were tested using the following methods:

- **Direct Saline Preparation**: A drop of saline was mixed with a small amount of stool on a glass slide and examined under a microscope for motile parasites and cysts.
- **Iodine Preparation**: A drop of iodine was added to a small amount of stool on a glass slide to stain cysts and ova, making them more visible under a microscope.
- **Sedimentation Concentration Techniques**: This involved mixing stool with saline, centrifuging the mixture, and examining the sediment for parasites.

### Data Analysis

The collected data were analyzed using IBM SPSS Statistics for Windows, version 21.0 (IBM Corp., Armonk, NY, USA). Descriptive statistics were used to summarize the data. The chi-square test was applied to determine the association between intestinal parasitic infections and various risk factors. Logistic regression analysis was performed to identify significant predictors of infection. A p-value of ≤ 0.05 was considered statistically significant.

## Results

### Demographics

Out of 201 primary school children, the majority were females (127, 63.2%), with a total median ± interquartile range (IQR) age of 10 ± 5 years. The minimum and maximum ages were 7 and 15 years, respectively.

**Table 1.**
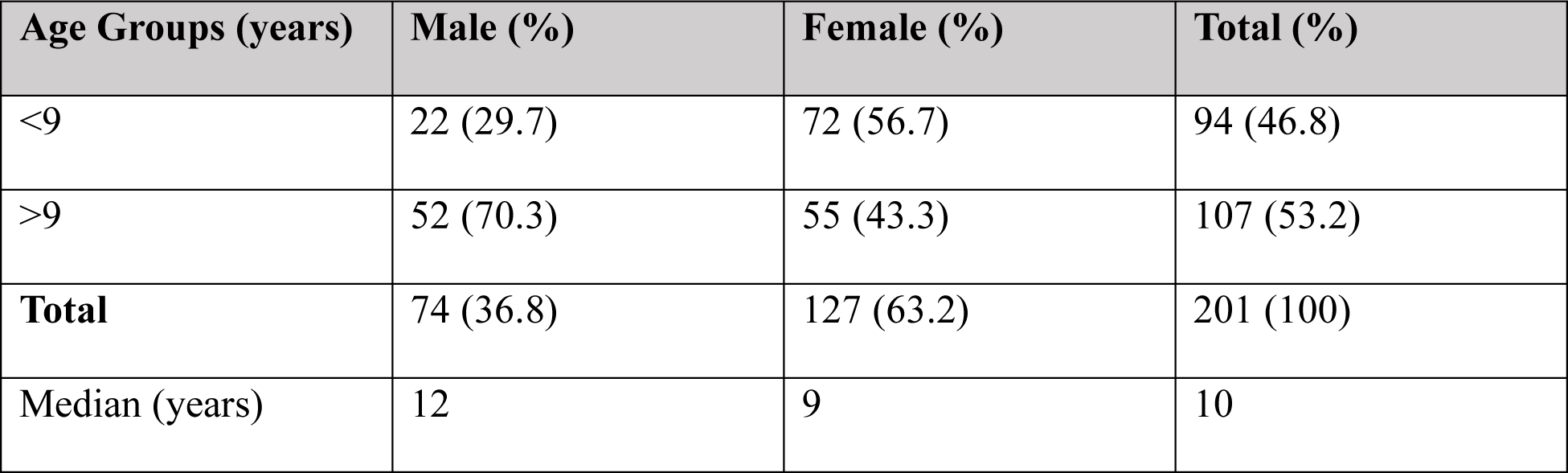
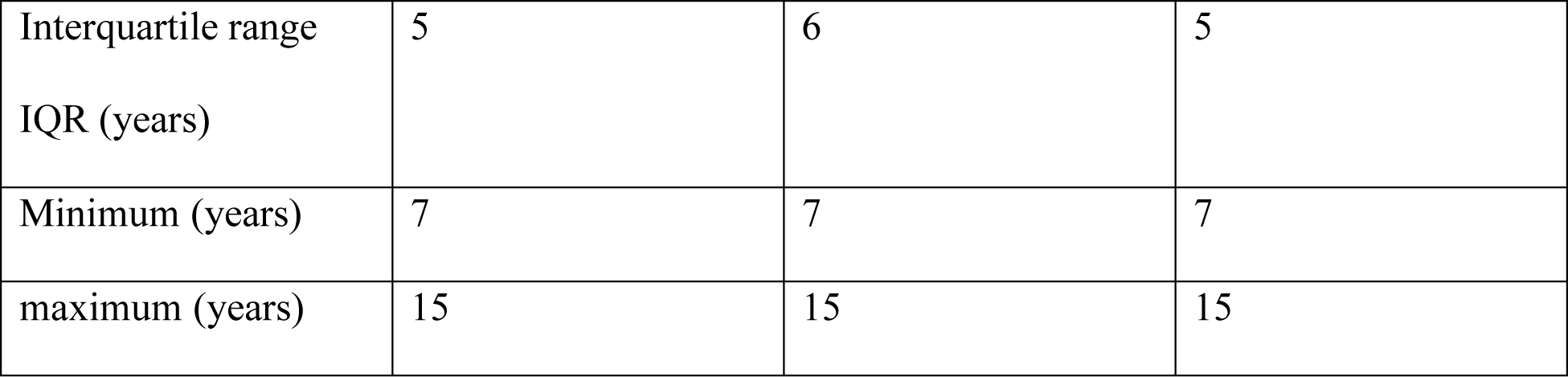
Age and sex distribution of schoolchildren in Aden, Yemen.

### Prevalence of Intestinal Parasites

The overall prevalence of intestinal parasites was 47.3%, with 35.8% having a single parasite and 11.5% having multiple parasites.

**Figure 1.**
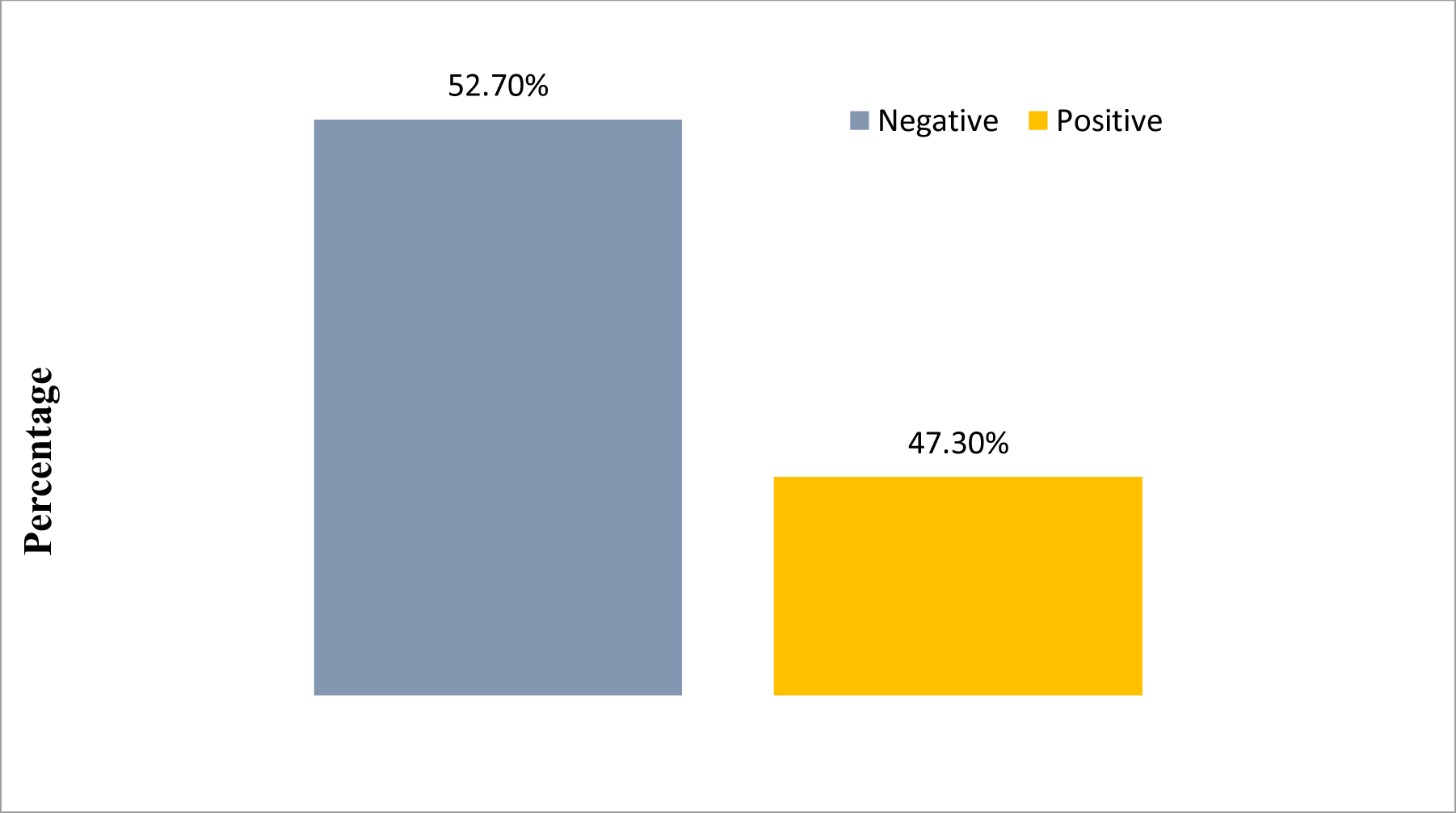
Prevalence of parasitic infection among primary schoolchildren in Aden, Yemen.

### Distribution of Intestinal Parasites by Sex

The prevalence rates were higher among female schoolchildren (51.2%) compared to males (45.5%).

### Distribution of Different Intestinal Parasites

The most predominant parasite identified was *Entamoeba histolytica/dispar*, found in 36.3% of the cases, followed by Giardia lamblia (15.4%), *Hymenolepis nana* (8.5%), and Enterobius vermicularis (5.0%).

### Single vs. Multiple Parasitic Infections

Among the children infected with intestinal parasites, 35.8% had a single parasitic infection, while 11.5% had multiple parasitic infections.

### Prevalence of Protozoa and Helminths

The prevalence of protozoa was 51.7%, while helminths were found in 8.9% of the infected children.

### Distribution of Intestinal Parasites by Maternal Education

The highest prevalence rates were observed among children whose mothers had primary school education (51.3%), followed by those whose mothers had secondary school education (50%).

### Distribution of Intestinal Parasites by Age Group

The prevalence of intestinal parasites was higher among children aged >9 years (55.1%) compared to those aged <9 years (50.0%).

### Distribution of Intestinal Parasites by Maternal Occupation

The prevalence rates were higher among children whose mothers were housewives (48.5%) compared to those whose mothers were employed (40.0%).

### Risk Factors for Intestinal Parasitic Infections

The analysis of risk factors for contracting intestinal parasitic infections among primary schoolchildren is shown below.

**Table 2.** Risk factors contracting for intestinal parasitic infections among primary schoolchildren in Aden, Yemen.

| Risk Factors | Number of Children with Positive Response (n) | Infection Positive (%) | P-value |
| --- | --- | --- | --- |
| Hand washing before meal | 188 | 90 (47.9%) | 0.511 |
| Hand washing after meal | 197 | 93 (47.2%) | 0.912 |
| Washing anal area with hand after defecation | 201 | 95 (47.3%) | Undefined |
| Nail hygienic status | 137 | 64 (46.7%) | 0.820 |

## Discussion

Our study found that the overall prevalence of intestinal parasitic infections among primary school children in Aden, Yemen, was 47.3%, with 35.8% having a single parasitic infection and 11.5% having multiple infections. The most predominant parasite identified was Entamoeba histolytica/dispar, present in 36.3% of cases. The prevalence was higher among female schoolchildren (51.2%) compared to males (45.5%). Additionally, the infection rates were highest among children whose mothers had primary school education (51.3%) and those whose mothers were housewives (48.5%). Children aged over 9 years had a higher prevalence rate of 50%. There was no significant association between the identified risk factors and intestinal parasitic infections.

Various studies have determined the prevalence of intestinal parasitic infections among primary schoolchildren globally. Taheri et al. found a similar rate of 47.7% in Iran [11]. A slightly similar finding was reported as 45.5% in Nigeria [12]. A study conducted in Saudi Arabia reported an overall prevalence of parasitic infection at 48% [13]. Another study performed in Yemen showed a prevalence of parasitic infection at 38.2% [5]. Forson et al. reported a rate of 15% in Ghana [14]. On the other hand, our result was much lower compared to other studies such as in Yemen, which reported prevalences of 51.26%, 57.4%, 61.85%, 66.3%, and 90% [6, 7, 15, 8, 16], North West Ethiopia at 84.3% [17], Iran at 68.1% [18], and Ethiopia at 51.5% [19]. The difference might be attributed to geographical differences, the living and socioeconomic nature of the study subjects, sample size, and the methods used for diagnosis [8].

The current study clearly showed that the prevalence of intestinal parasitic infection was higher in females (51.2%) than males (45.5%) (Figure 2). Simon-Oke et al. also reported that females had more parasitic infections than males [20]. Other studies conducted in Nepal, Morocco, Saudi Arabia, Ethiopia, and Yemen reported higher infections among females [21, 13, 22, 23, 8].

**Figure 2.**
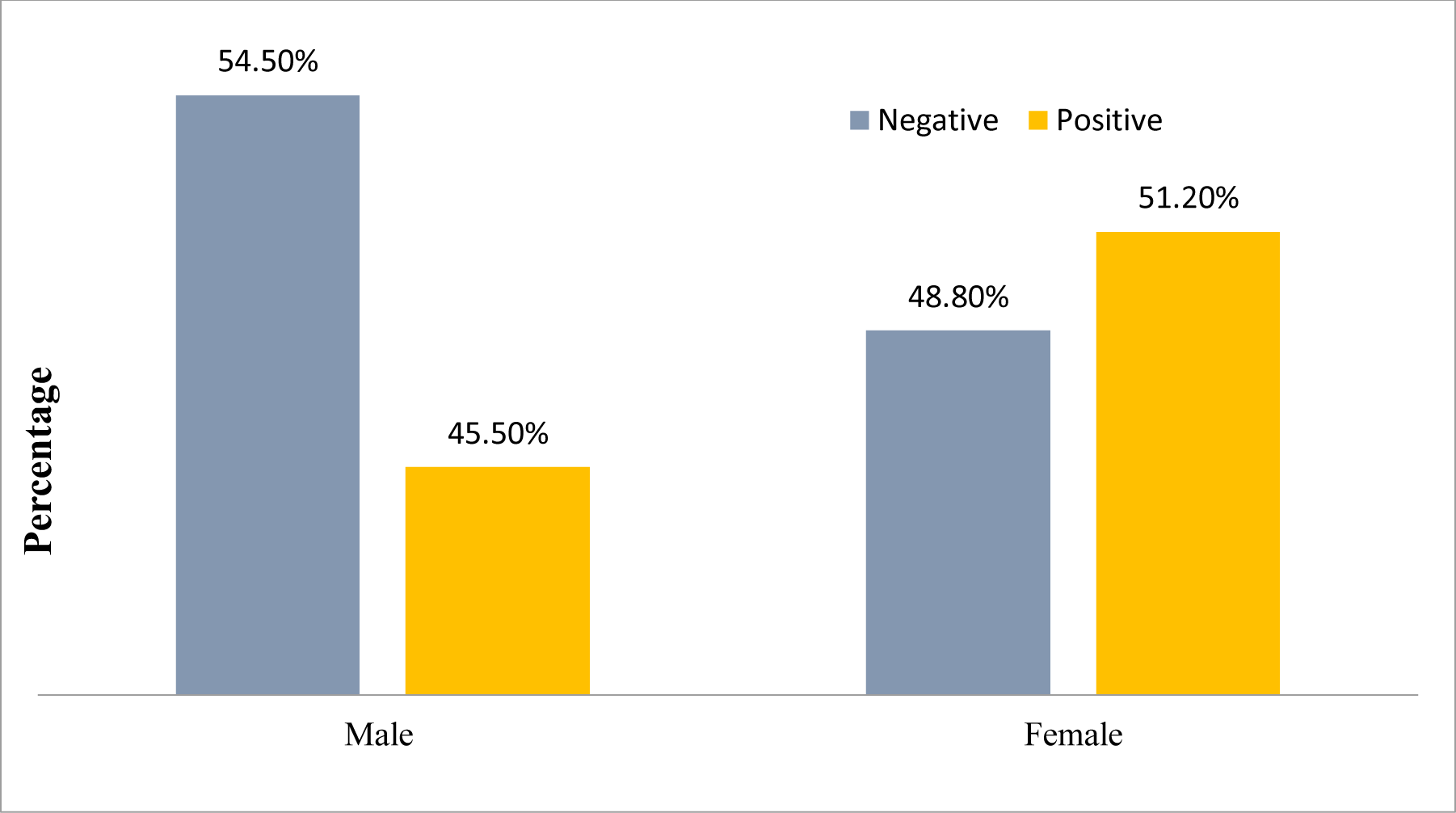
Prevalence of parasitic infection among primary schoolchildren according to their sex in Aden, Yemen.

Conversely, other studies have indicated an opposite finding where the rate was higher among males, as in Yemen [15, 16], Ethiopia [24, 25], Nigeria [26], and Northwest Ethiopia [27]. This indicates that sex may play a role in parasitosis depending on the region and other environmental or behavioral factors [28].

The findings of this analytical cross-sectional data showed that E. histolytica/dispar was the most predominant parasite among schoolchildren (36.3%) (Figure 3). Similar findings were shown in Yemen and Burkina Faso [7, 29, 15, 16]. Lone et al. recorded that the most common detected parasite was Ascaris lumbricoides in India [30]. Other studies conducted in Ghana, Sudan, Nepal, Yemen, and Iran revealed that G. lamblia was the most frequent protozoan infection [14, 31, 32, 8, 11]. A study done in Ethiopia reported that the most prevalent intestinal parasite was hookworm [17]. Entamoeba coli was identified as the most common parasite in a study undertaken in Egypt [33]. Gelaw et al. found that Hymenolepis nana was the most predominant parasite in Ethiopia [27]. Hailegebriel reported that Entamoeba histolytica/dispar and hookworm were common parasites among children in Ethiopia [34]. Several factors may determine the distribution of parasitic infection among children, including personal hygiene, water sources, economic status, and parental occupations [32].

**Figure 3.**
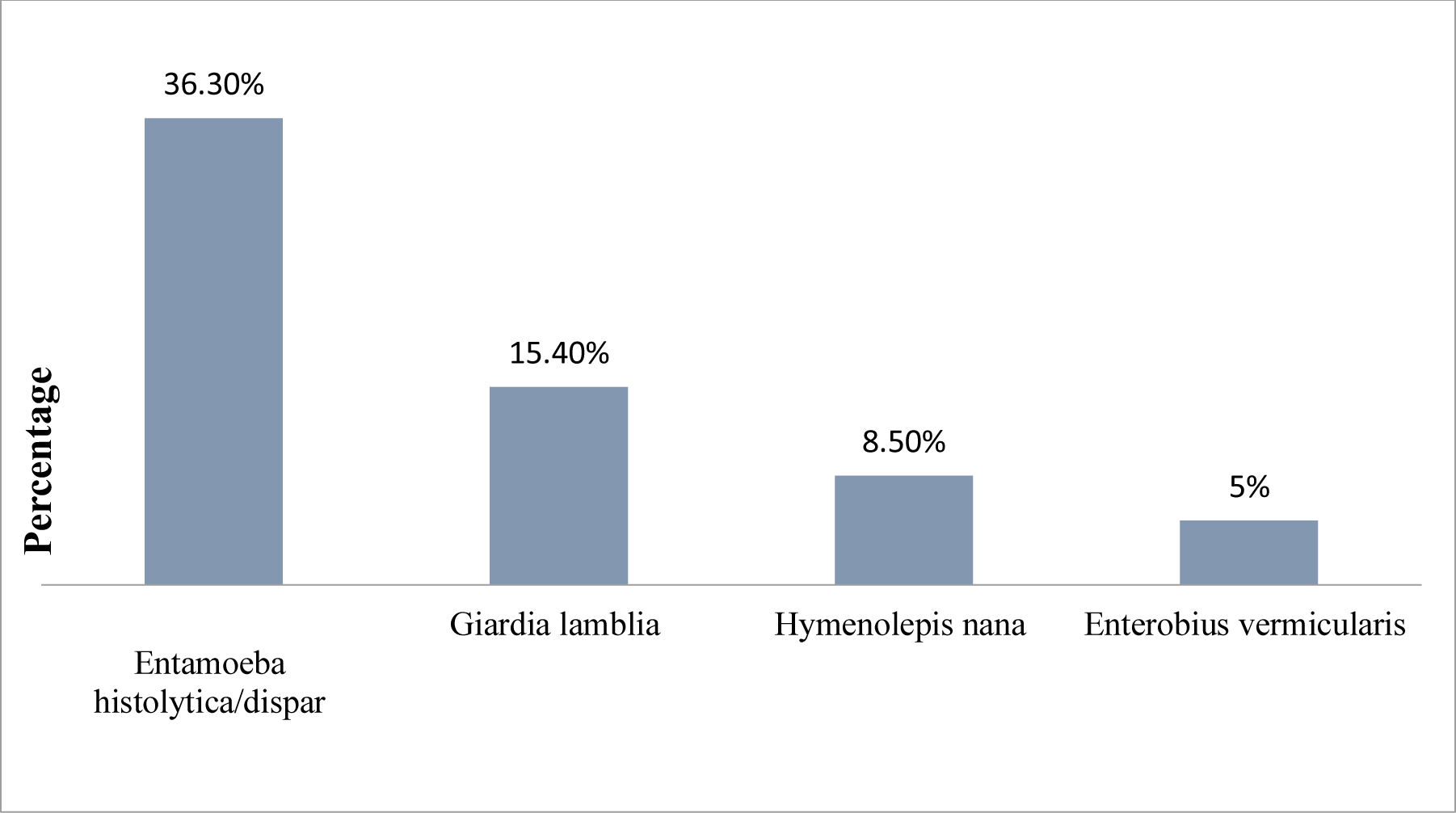
Distribution of different intestinal parasites detected among primary schoolchildren in Aden, Yemen.

Out of the 47.3% children positive for intestinal parasites, 35.8% had one parasite, and 11.5% had multiple parasitic infections (Figure 4). A result from Ghana showed that a single parasite was found among 13.6% of students while double infections were among 1.3% of students [14]. A study performed in Ethiopia reported that multiple infections occurred in 13.5% of schoolchildren [27]. Edrees et al. showed that 23.5% of children had multiple infections in Yemen [16].

**Figure 4.**
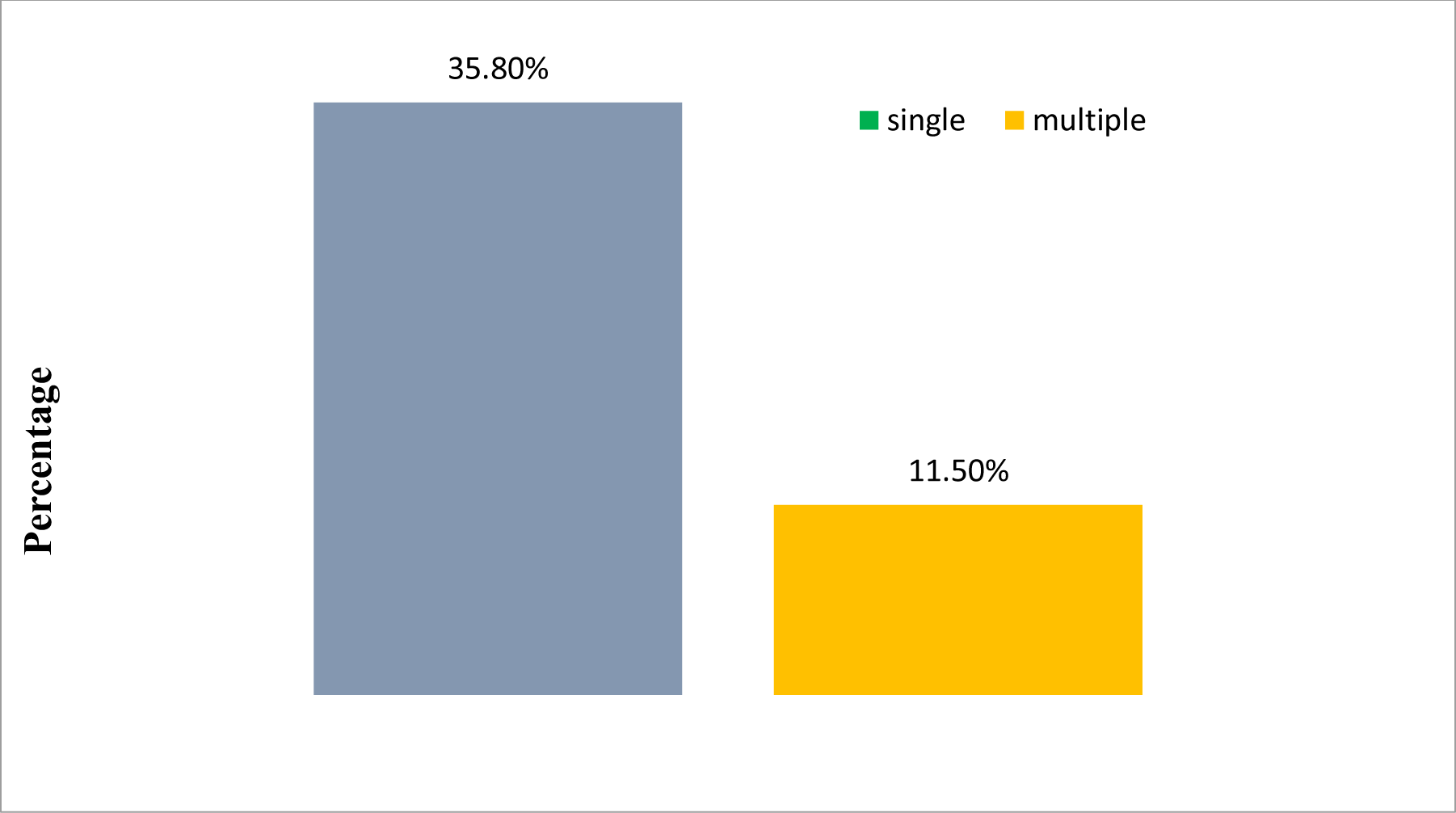
Presence of single or multiple parasitic infections among primary schoolchildren in Aden, Yemen.

In the current study, protozoan infections were higher than helminths infections (Figure 5). Erismann et al., Gupta et al., and Edrees et al. revealed the same result in Burkina Faso, India, and Yemen, respectively [29, 32, 16]. Gelaw et al. found that helminthes were higher than protozoan infections in Ethiopia [27]. The reason for the difference might be the geography of the place, socioeconomic conditions of the study area, and the hygienic habits of the studied group [35].

**Figure 5.**
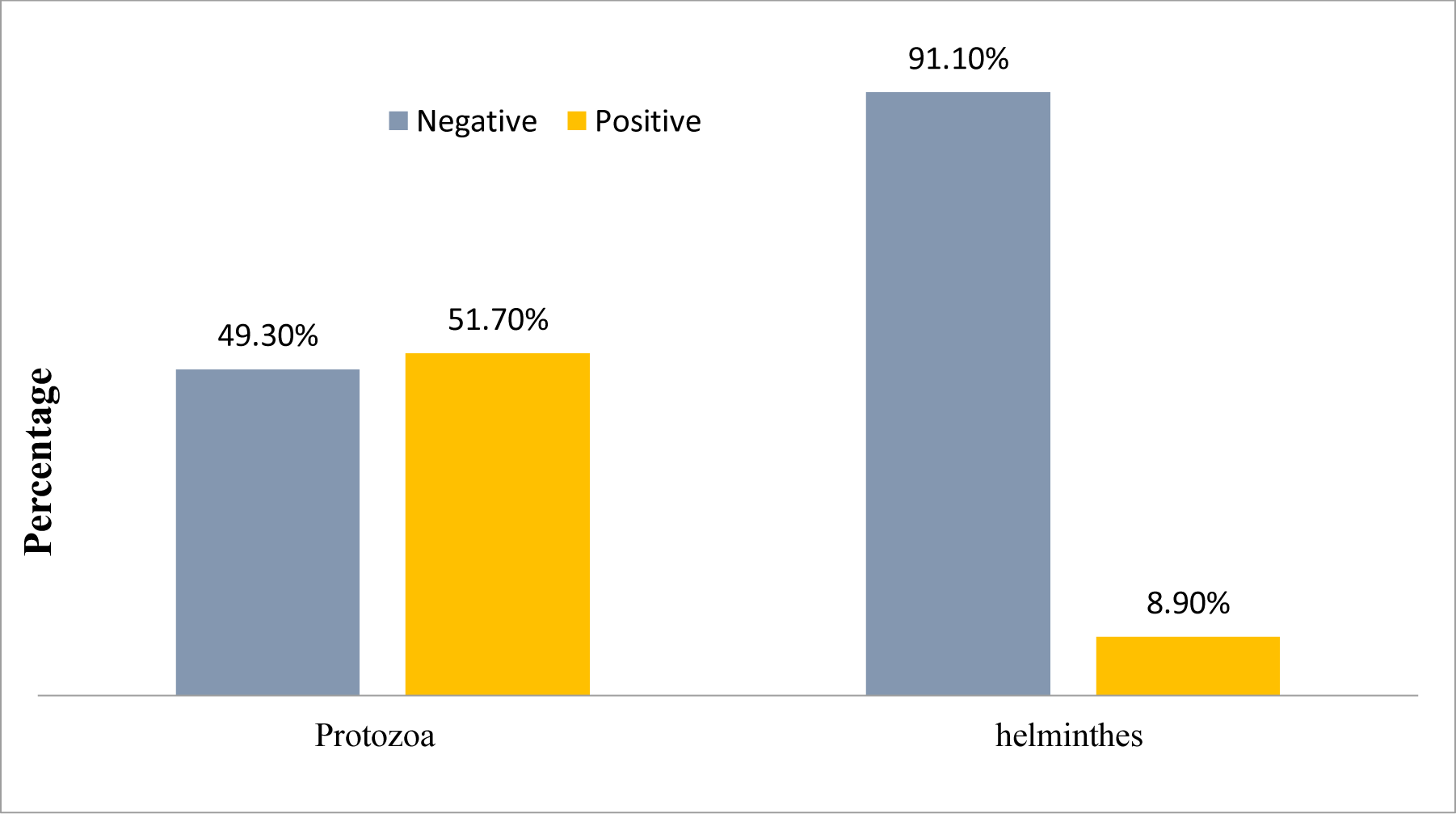
Prevalence of protozoa and helminths among primary schoolchildren in Aden, Yemen.

This study’s results showed the highest prevalence rate of parasitic infection was 51.3% among children whose mothers had primary school education, followed by 50% among secondary school educated mothers (Figure 6). Alemu et al. and Sitotaw & Shiferaw showed that the highest rates were among children whose mothers had primary school education in Ethiopia [24, 23]. Our data disagreed with a study done in Jabel Aweleia Governorate, Khartoum, which recorded the highest percentage of infected children whose mothers attended Quran school, followed by those whose mothers were illiterate [36]. Hailegebriel reported that illiterate mothers were associated with high infection rates among their children in Ethiopia [34]. According to maternal occupation in the current data, the high prevalence was 48.5% among children whose mothers were housewives (Figure 7). This was in agreement with studies carried out in Khartoum and Ethiopia [36, 23]. On the other hand, a study conducted in rural Khartoum found the highest percentage among children whose mothers were unskilled laborers [36]. Differences may arise from levels of awareness and education among housewife mothers in various countries. In contrast, the workload of employed mothers, leading to spending valuable time away from their children, was a risk factor that increased parasitic infections among young children [37].

**Figure 6.**
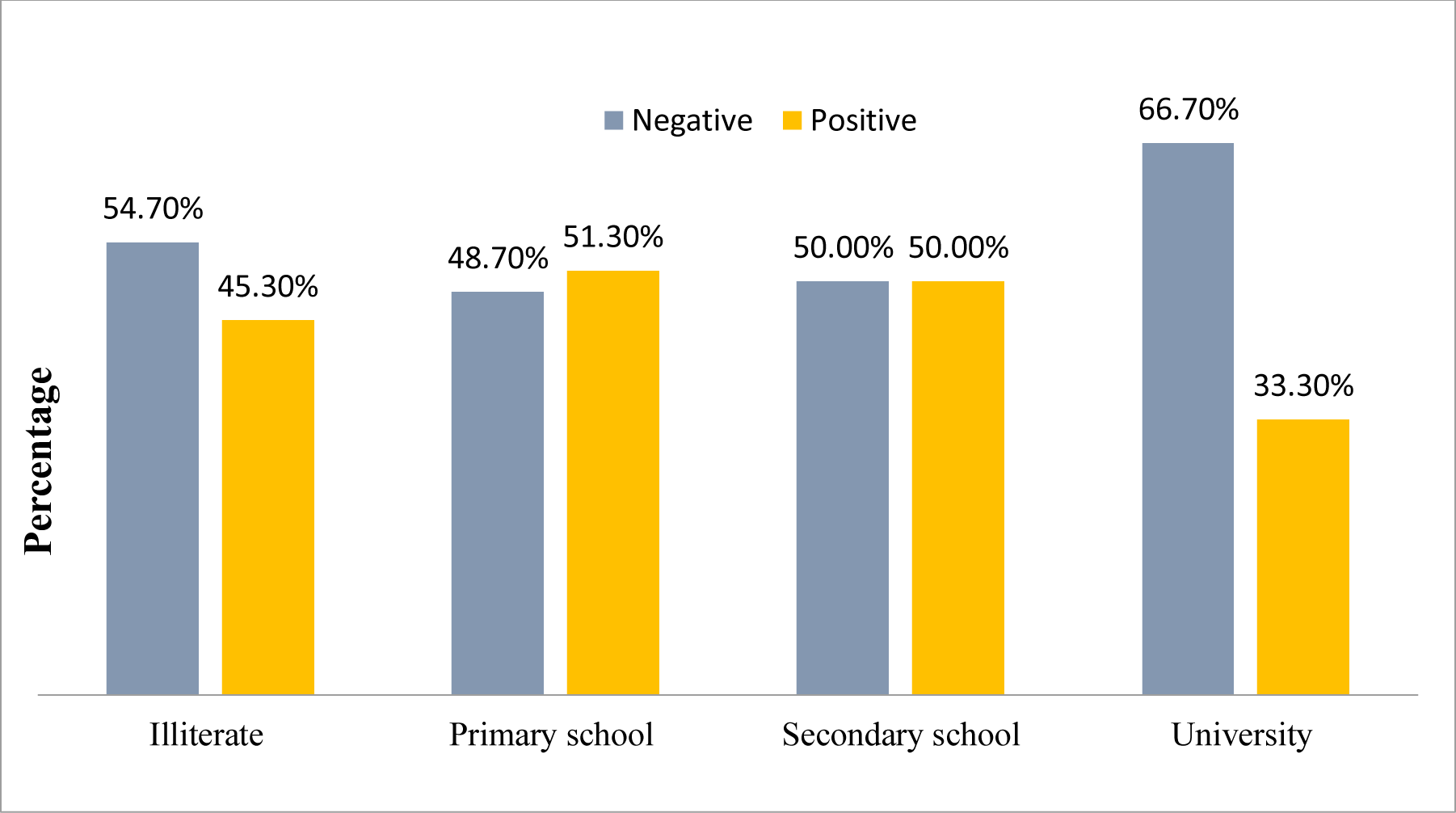
Prevalence of parasitic infection among primary schoolchildren according to the education of their mother in Aden, Yemen.

**Figure 7.**
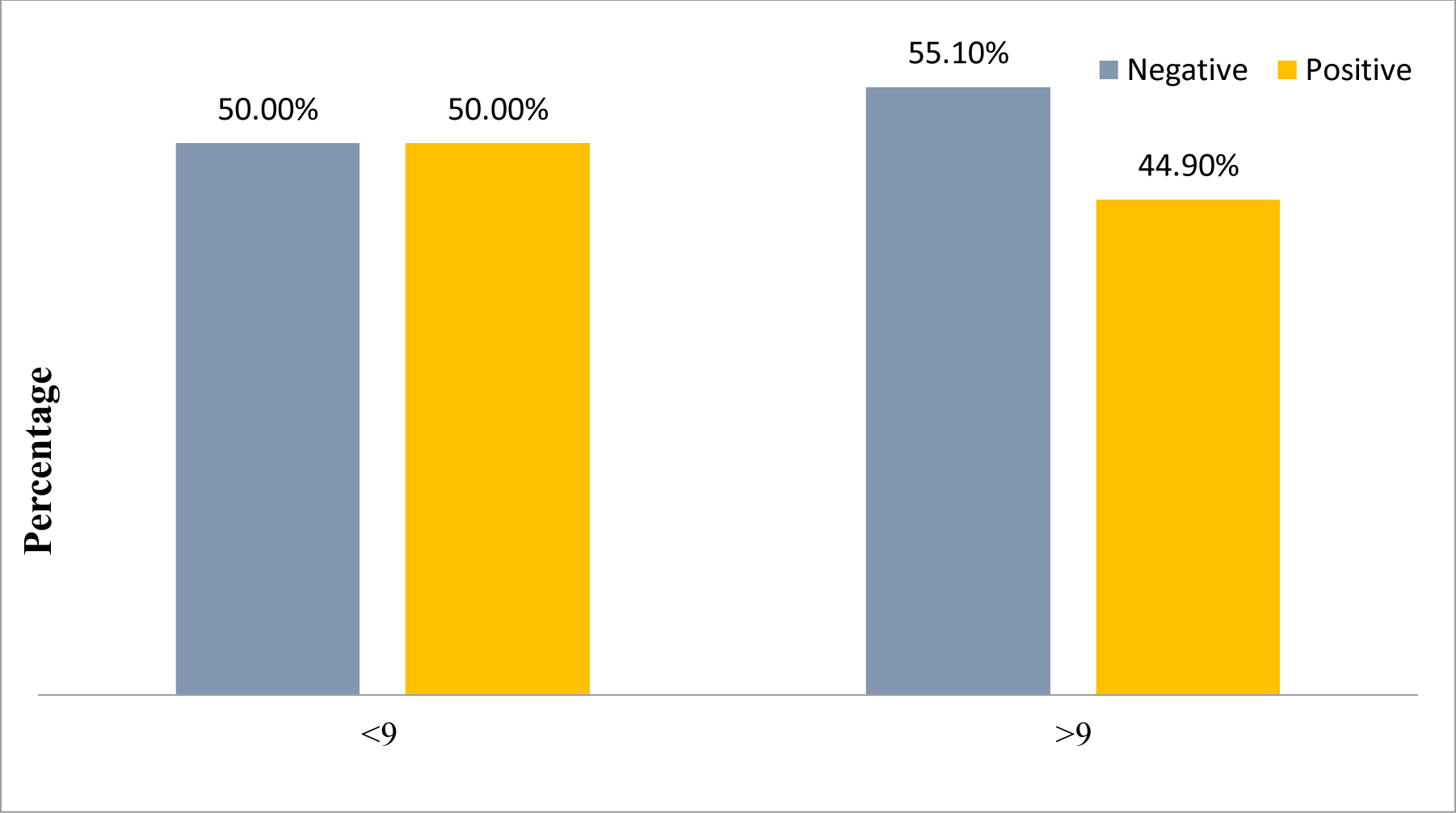
Prevalence of parasitic infection among primary schoolchildren according to their age in Aden, Yemen.

The results of this study clearly showed that the prevalence of infection was high (50%) in the age group >9 years (Figure 8). This agreed with studies from Yemen [7, 6]. Sitotaw et al. detected the highest rate in the age group 2–18 years in Ethiopia [24]. This disagreed with that reported in Burkina Faso [29]. A study conducted in Morocco showed that the infection rate was highest among children aged more than 10 years [22]. A study conducted in Nepal showed the highest rate among children aged less than 6 years [38]. This may be because children in this age group do not pay much attention to their personal hygiene, wearing protective shoes, and sharing food using dirty or unclean hands, which exposes them to parasitic infection [3].

**Figure 8.**
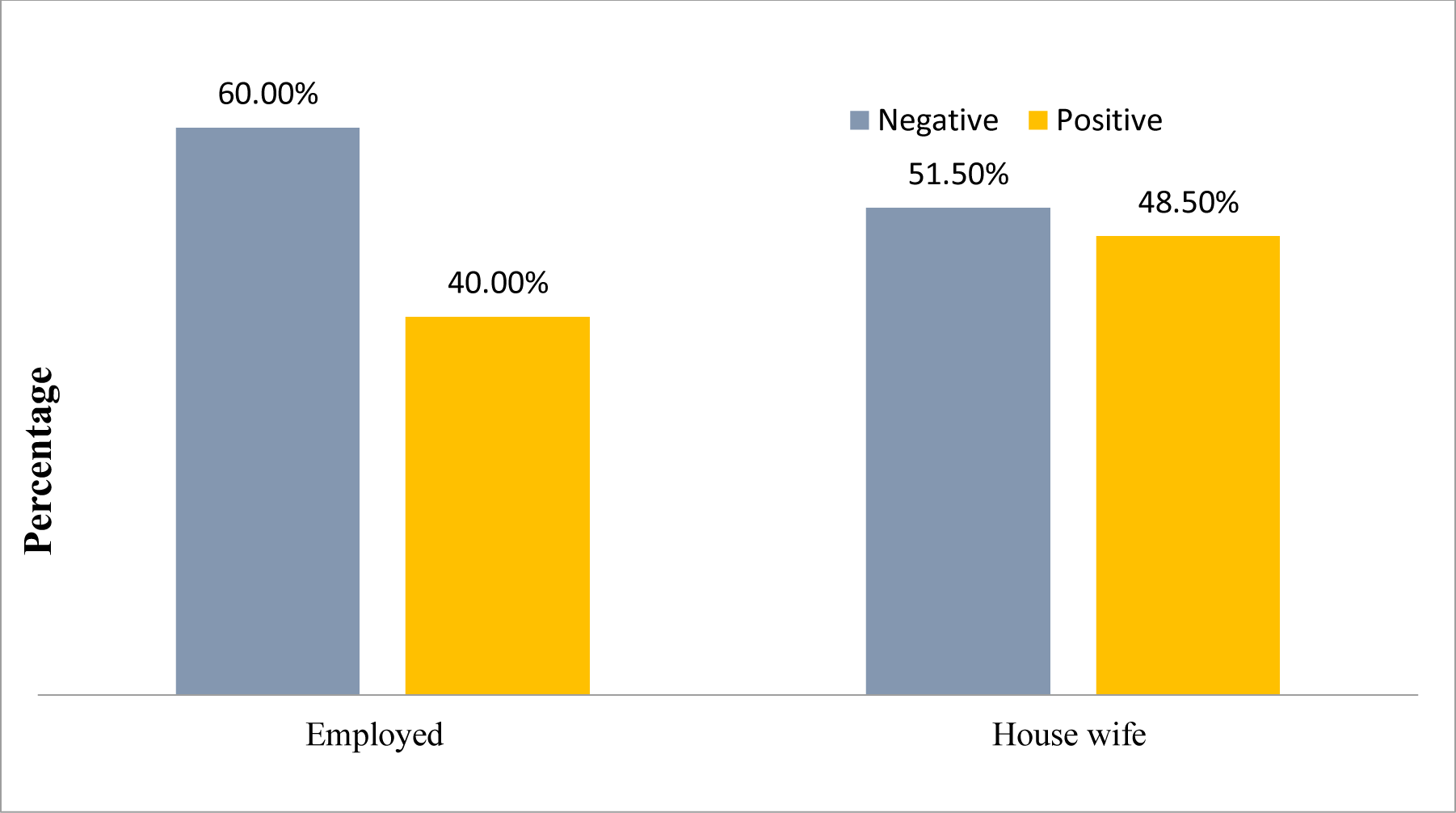
Prevalence of parasitic infection among primary schoolchildren according to occupation of their mothers in Aden,Yemen.

The main limitation of this study was that the identification of some parasites may have been missed due to difficulty in using suitable techniques such as cellophane tape for Enterobius vermicularis or modified acid-fast stain for intestinal coccidian.

## Conclusions

The prevalence rate of intestinal parasites was found to be higher in Aden, Yemen. The most dominant parasite was Entamoeba histolytica/dispar, with the highest rates among female schoolchildren, children whose mothers were housewives, those with primary and secondary school education, and children aged >9 years.

## Data Availability Statement

All relevant data are within the paper.

## Author Contributions

- **Conceptualization**: Ali N. M. Gubran, Naif Mohammed Al-Haidary
- **Methodology**: Ali N. M. Gubran
- **Investigation**: Marwa Faisal M. Bajubair, Afrah Mohsen Ali Algibary, Manal Galeb Mhmad Ali, Marwa Fuad Othman Ali
- **Data Curation**: Marwa Faisal M. Bajubair, Afrah Mohsen Ali Algibary, Manal Galeb Mhmad Ali, Marwa Fuad Othman Ali
- **Formal Analysis**: Ali N. M. Gubran, Naif Mohammed Al-Haidary
- **Writing - Original Draft**: Ali N. M. Gubran, Naif Mohammed Al-Haidary
- **Writing - Review & Editing**: Ali N. M. Gubran, Naif Mohammed Al-Haidary
- **Supervision**: Ali N. M. Gubran, Naif Mohammed Al-Haidary

All authors have reviewed and agreed on their contributions to this manuscript.

